# Protocol for the Tessa Jowell BRAIN MATRIX Platform Study

**DOI:** 10.1101/2022.07.29.22277991

**Authors:** Colin Watts, Joshua Savage, Amit Patel, Rhys Mant, Victoria Wykes, Ute Pohl, Helen Bulbeck, John Apps, Rowena Sharpe, Gerard Thompson, Adam Waldman, Olaf Ansorge, Lucinda Billingham, the TJBM Investigators

## Abstract

**Introduction:** Gliomas are the most common primary tumour of the central nervous system (CNS), with an estimated annual incidence of 6.6 per 100,000 individuals in the USA and around 14 deaths per day from brain tumours in the UK. The genomic and biological landscape of brain tumours has been increasingly defined and, since 2016, the WHO classification of tumours of the CNS incorporates molecular data, along with morphology, to define tumour subtypes more accurately. The Tessa Jowell BRAIN MATRIX Platform (TJBM) study aims to create a transformative clinical research infrastructure that leverages UK NHS resources to support research that is patient centric and attractive to both academic and commercial investors.

**Methods and analysis:** The TJBM study is a programme of work with the principal purpose to improve the knowledge of glioma and treatment for glioma patients. The programme includes a platform study and subsequent interventional clinical trials (as separate protocols). The platform study described here is the backbone data-repository of disease, treatment and outcome data from clinical, imaging and pathology data being collected in glioma patients from secondary care hospitals. The primary outcome measure of the platform is time from biopsy to integrated histological-molecular diagnosis using whole genome sequencing and epigenomic classification. Secondary outcome measures include those that are process-centred, patient-centred and framework-based. Target recruitment for the study is 1000 patients with interim analyses at 100 and 500 patients.

**Ethics and dissemination:** The protocol was approved by West Midlands - Edgbaston Research Ethics Committee. Participants will be required to provide written informed consent. The results of this study will be disseminated through national and international presentations and peer-reviewed publications.

**Trial registration number:** ClinicalTrials.gov Identifier: NCT04274283; 18-Feb-2020

ISRCTN 14218060; 03-Feb-2020

**Strengths and limitations of this study:**

- The TJBM study is a programme of work, which will improve the knowledge of glioma, and treatment for glioma patients.
- This platform study is a backbone data-repository of disease, treatment and outcome data from clinical, imaging and pathology data
- The TJBM study will aim to provide rapid and accurate molecular diagnosis, a network of clinical hubs with robust protocols for the collection, processing, analysis and storage of tissue, images, and clinical and quality of life data
- Gliomas occur at all ages and their specific subtype is difficult to predict preoperatively, therefore, the patient population eligible for the study is broad but currently excludes <16-year-old patients

## Introduction

Gliomas are the most common primary tumour of the central nervous system (CNS), with an estimated annual incidence of 6.6 per 100,000 individuals in the USA (1), which is predicted to rise to 22/100,000 by 2035 (2). In 2016, there were 5,250 deaths from brain tumours in the UK i.e., 14 deaths per day (2). Malignant CNS tumours hold the poorest prognosis, and are responsible for the highest estimated number of years of potential life lost (mean 20 years) amongst all cancers (3) and survival trends have remained generally static in comparison to other cancers (4).

Gliomas have traditionally been divided into low grade glioma (LGG; WHO I-II) and high-grade glioma (HGG; WHO III-IV) based on integrated classic histological features and molecular biomarkers (5). LGG have an indolent course with patients commonly surviving a decade after diagnosis (6). However, the natural history of many WHO Grade II LGG is progression to HGG. Approximately half of all newly diagnosed gliomas are classified as glioblastoma (GB; WHO IV), the most malignant type of brain cancer. Currently, the GB annual incidence is 3.2 per 100,000 population in the USA. This tumour occurs more frequently with advancing age; ranging from 0.4 per 100,000 population aged 20–34 years, to over 15 per 100,000 population aged 75-84 years (1). It is widely recognized that elderly populations are rapidly increasing globally and this will have a significant impact on the burden of GB disease. Despite this, most studies still focus on patients younger than 65 years.

The current gold standard treatment for newly diagnosed GB is surgical gross total resection (GTR), followed by radiotherapy with concomitant and adjuvant temozolomide. The aim of treatment is to delay tumour progression and extend overall survival (7). Despite decades of refinement, this approach results in a median survival time of only 12-14 months.

Over the last 20 years the genomic and biological landscape of brain tumours has been increasingly defined, refining previous classification systems, unravelling intra- and inter-tumoural heterogeneity and progression, identifying drug targets and potential therapeutic strategies, and better characterising challenges to therapies, such as the blood brain barrier (BBB) and mutational escape. In 2016, the WHO published its classification of tumours of the CNS (5), which for the first time used molecular data, along with morphology, to define tumour subtypes more accurately. This stratification integrated a combination of specific genetic and epigenetic biological characteristics that are changing rapidly as our knowledge evolves (8). In recognition of this rapid evolution of knowledge, ‘cIMPACT-NOW’ (Consortium to Inform Molecular and Practical Approaches to CNS Tumor Taxonomy – Not Official WHO) (9) was created, with the new WHO 2021 classification representing a consensus opinion based on new insights into the molecular definition of brain tumours from this collaboration (10). As analytical technologies evolve and become more affordable, genome-wide analyses combined with other so-called ‘omics’ data will lead to further refinement of tumour classification with meaningful impacts on prognosis and choice of therapy. This is the motivation for the TJBM study, which will adopt whole-genome sequencing (WGS) and epigenomic analysis of tumour samples.

The TJBM study aims to create a transformative clinical research infrastructure that leverages NHS resources to support research that is patient centric and attractive to both academic and commercial investors. Our approach will be holistic and flexible so that it can rapidly adapt to scientific advances and rigorously evaluate new drugs and technologies. To achieve this, it will establish a research-active network of clinicians and scientists who are well supported by an innovative trial infrastructure to deliver the following:

- Rapid and accurate molecular diagnosis ensuring precise classification of tumours and identifying subsets of patients suitable for targeted therapy, by building on the legacy of the 100,000 Genomes Project (11).
- A network of clinical hubs resourced to maximise patient recruitment and collect tissue and data.
- Robust protocols for the collection, processing, analysis and storage of tissue, images and clinical and quality of life (QoL) data, building on existing infrastructure such as UK Biobank (12), Medical Research Council (MRC) Brain Banks Network (13), and the CRUK PEACE study (14).
- High-quality biological samples that are fully clinically and radiologically annotated facilitating further biological and radiological research.
- Links to national and international clinical and scientific infrastructure and networks (15); e.g., Genomics England (GEL), European Network for Rare adult solid Cancer (EURACAN) (16), SIOP-E Brain Tumour Group, and National Cancer Research Institute (NCRI) Groups.
- Access to novel and repurposed drugs and technologies through collaborative partnerships with industry and early phase trials hubs and the structural genomics consortium (www.thesgc.org), to develop novel trials, within and outside, the TJBM infrastructure for testing of therapeutic strategies, including novel agents. As such therapeutic clinical trials will be developed using this infrastructure to maximise efficiency in introducing and rigorously evaluating novel interventions. These will be separate protocols to the TJBM study.
- Long-term sustainability through delivery of clinically and scientifically meaningful outcomes, leveraged investigator-led research grants and an established cost-recovery model for biobanking (17).

## Methods and analysis

### Study design

The TJBM study is a programme of work the principal purpose of which is to improve the knowledge of glioma and treatment for glioma patients. The programme will include a platform study and subsequent interventional clinical trials (as separate protocols); Figure 1. This platform study, is the backbone data-repository of disease, treatment and outcome data from clinical, imaging and pathology data to be collected in glioma patients from secondary care hospitals. Figure 2 shows an overview of the platform’s study schema.

**Figure 1:**
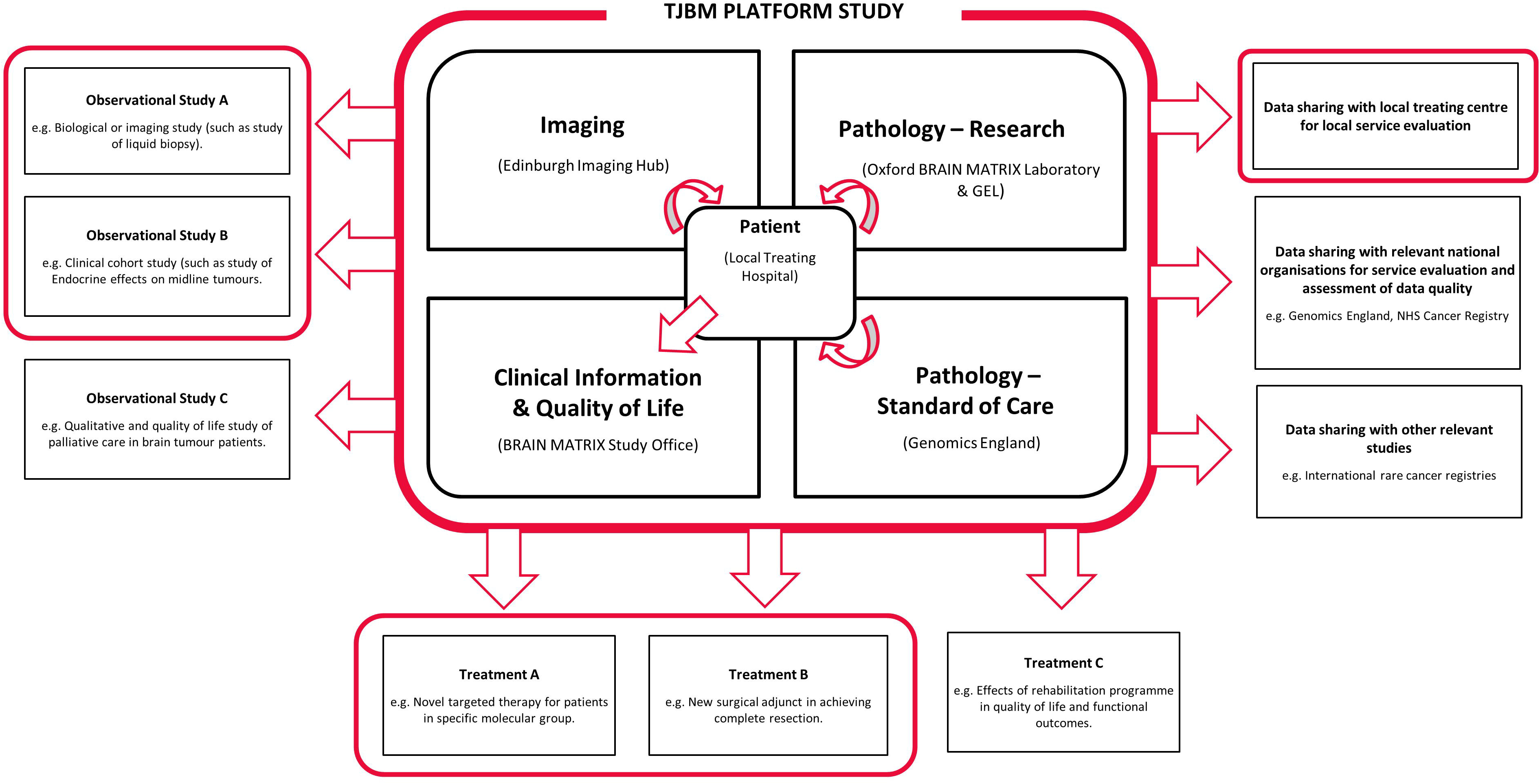
Overview of the Tessa Jowell BRAIN MATRIX Programme. The Tessa Jowell BRAIN MATRIX Platform (TJBM) study will collect and integrate clinical, pathological, advanced molecular, imaging, quality of life, treatment and outcome data. The platform may provide data directly or support identification of eligible patients to clinical trials, within and outside the TJBM study programme. If eligible, patients may be enrolled in multiple add-on studies. Through consent and with strong governance processes, anonymised or pseudonymised data may be shared with other relevant organisations or studies within and outside the programme.

**Figure 2:**
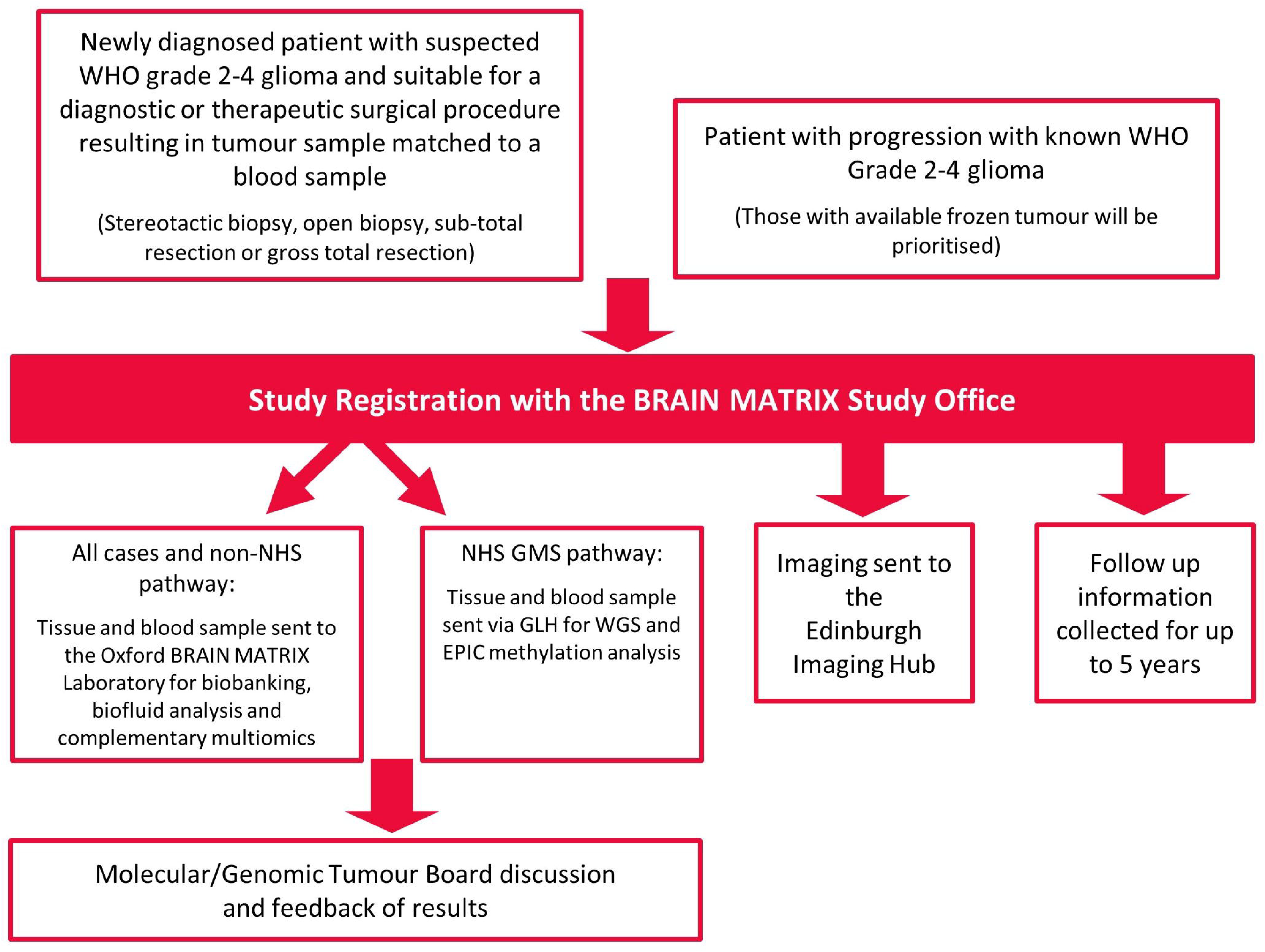
The Tessa Jowell BRAIN MATRIX Platform study schema. The study schema for the Tessa Jowell BRAIN MATRIX Platform study.

The study aims to recruit 1000 patients within the UK over 5 years with participants followed up for up to 5 years. An initial 10 UK centres were opened to the TJBM study, as listed in Supplementary Appendix 1, although further centre expansion is planned. The Standard Protocol Items: Recommendations for Intervention Trials (SPIRIT) checklist is provided as Supplementary Appendix 2 (18). The World Health Organization (WHO) Trial Registration Data Set is provided in Supplementary Appendix 3.

### Patient and public involvement

Patient and public involvement and engagement (PPIE) has been integral to this study from its inception. Our patient and public advisors, Helen Bulbeck and Peter Buckle, co-developed the TJBM study by reviewing and refining the protocol and the participant-facing documents. They have provided input into the patient-reported outcome measures and have guided messaging about the study for the community. As members of the Study Management Group (SMG; Supplementary Appendix 1) they continue to assess study conduct, and will contribute to the interpretation and dissemination of study findings through the PPIE dissemination strategy. This will include presentation of the study findings to identified audiences, the best channel for reach, when to communicate, messaging and sensitivities.

### Patient selection

Patients aged ≥16 years with newly diagnosed suspected WHO Grade II-IV glioma, (as evidenced radiologically) suitable for a diagnostic or therapeutic surgical procedure resulting in a tumour sample matched to a blood sample or with progression with known WHO Grade II-IV glioma will be eligible.

Patients with primary spinal cord tumours, who are receiving active treatment of other malignancy, have contraindications to magnetic resonance imaging (MRI) and/or without standard of care imaging available, will be excluded.

Newly diagnosed patients with suspected WHO Grade II-IV glioma who are subsequently found to have a WHO Grade I tumour or non-brain tumour are expected to be a rare occurrence. If this event occurs:

- If it is confirmed as a Grade I tumour, then the patient will remain eligible for the study and will continue to be followed up in accordance with the protocol.
- If it is confirmed as a non-brain tumour, then the patient will not remain on the study and no further follow-up data will be collected.

All patients must be able to provide written informed consent for the study.

### Consent

Supplementary Appendix 4 contains exemplar informed consent forms (ICF), with Supplementary Appendix 5 the patient information sheets (PIS) and lay summary for the study. The investigator or an appropriately trained delegate must obtain written informed consent for each patient prior to performing any study related procedure. Remote Consent for platform entry is permitted by telephone or video consultation instead of face-to-face consultations.

In addition, to consenting to join TJBM, patients will be required to provide agreement for:

- The collection and analysis of biological samples (e.g., tumour, blood), including access to existing and future samples.
- The collection of relevant clinical information, including imaging and pathology.
- The return of clinically relevant results back to the referring clinician.
- The use of, and sharing of data, for research, teaching, commercial and scientific purposes, including data sharing through The Brain Tumour Charity’s BRIAN (the Brain tumouR Information and Analysis Network) database (19).
- The collection of different aspects of health data from the NHS and other Department of Health organisations (in addition to medical records) for longitudinal analysis and follow-up.
- The use of clinical data to identify potential clinical trials or other research that they may benefit from.
- The sharing of samples for other ethically approved research projects.

In addition, and in line with GEL processes and studies such as the 100,000 Genomes Project, optional consent will be taken as to whether the patient would like to receive feedback about the evidence of inherited diseases (both underlying the cause of the cancer and non-cancer causes).

For centres submitting tumour tissue for WGS through the standard of care (SoC) NHS Genomic Medicine Service (GMS) pathway in England, an additional GEL consent step is currently required to confirm consent at the point of referral for the patient’s clinical indication. This additional GEL consent step is performed electronically or on paper and can be completed over the phone.

### Platform assessments and schedule of events

Platform entry requires baseline clinical data, NIH Stroke Scale (20), weight, and WHO Performance Status to be recorded, and then performed again at start and end of concomitant therapy, start and end of adjuvant therapy, further surgery and during the five-year follow-up period.

In addition, QoL questionnaires will be completed at the initiation of concomitant therapy, initiation of adjuvant therapy, further surgery and during follow-up. The QoL booklet includes EQ-5D-5L (21), EORTC-QLQ-c30 (22), Patient Concerns Inventory (PCI) (23), Patient Global Impression of Change (PGIC) (24), and Clinician Global Impression of Change (25).

The schedule of events in included in Supplementary Appendix 6.

### Platform study outcome measures

The primary outcome measure of the platform is time from biopsy to integrated histological–molecular diagnosis (TTMD) as defined as the difference (in days) between date of biopsy and date of WGS and epigenomic classification.

Secondary outcome measures include those that are process-centred, patient-centred, and framework-based.

Process-centred secondaries include: time to completion of each node of tissue and imaging pathway; tumour and biological sample(s) quality control (QC) status; imaging QC status, and; inter-rater agreement of response assessment in neuro-oncology (RANO) (26).

Patient-centred secondaries include: extent of surgical resection; overall survival time (OS); intracranial progression-free survival time (PFS); QoL scores; type of interventions received; type of complications from treatments (standard of care) received, and; concordance between initial local radiological diagnosis, local pathological diagnosis, and integrated histological-molecular diagnosis.

Research framework-based secondaries include: samples and images centrally stored; targetable mutation(s) identified; post-mortem sampling consent status and sample collection confirmation, and; number of applications to, and outputs resulting from data repository (including trial proposals both within and outside of the TJBM network).

### Statistical analysis plan

The target sample size for the study is 1000 patients. The primary remit of TJBM study is to establish a central data repository that will support the development and delivery of precision medicine for all glioma patients in the UK. As such, there is no statistical basis behind the choice of target sample size, but this number will allow robust assessment of feasibility and subgroup analyses. Sample size for any clinical trials that are subsequently linked to the platform will be based on statistical justification.

A formal interim analysis of the primary outcome measure (TTMD) and any relevant secondaries will be performed after registration and diagnosis of the first 100 patients and after 500 patients. There are no formal stopping rules. Formal analyses of all study outcome measures will be performed once the study has completed recruitment (target 1000 patients) and completed the follow up for all registered patients.

The analysis of TTMD will essentially be descriptive. The median TTMD will be reported overall and for each centre, together with the proportion of patients achieving TTMD within 28 days. Graphs of the change in both these summary statistics over time will be used to explore if TTMD changes during the course of the study. All estimates will be accompanied by 95% confidence intervals (CI). The time to completion of each node of tissue and imaging pathway will be reported as medians together with 95% CI, both overall and for each centre. Swimmer plots will be used to depict overall and node level timings for each patient.

For each type of tumour and biological sample, the proportion of sample passing QC and successfully undergoing WGS and EC will be reported with 95% CI and similarly for the imaging QC outcome measures. Inter-rater agreement of RANO will be assessed through Kendall’s coefficient of concordance.

Extent of surgical resection is evaluated from the post-operative MRI scan and is categorised as either; closed biopsy, open biopsy, debulking <50%, subtotal resection 50-90%, near total resection 90-<100%, gross total resection 100%. The extent of surgical resection will be reported as the proportion falling into each category together with 95% CI.

OS is defined as the time from date of diagnosis to the date of death with patients who are alive at the time of analysis censored at the date last seen in clinic. Intracranial PFS time is defined as the time from date of registration to the earliest of date of intracranial progressive disease or death from disease. The date of an event is defined as the earliest confirmation of progression by radiological assessment, clinical symptoms or multidisciplinary team (MDT). Patients without progression at the time of analysis will be censored at the date last seen in clinic.

OS and intracranial PFS will be analysed and plotted using the Kaplan-Meier method. Median times with corresponding 95% CI will be reported together with rates at 1, 2 and 5 years. Multivariable survival regression modelling will be used to explore prognostic factors, including, but not limited to, WHO 2021 classification, age, tumour volume, stage, methylation and mutation status.

Longitudinal measures of QoL will be generated from the QoL questionnaire according to the questionnaire-specific algorithms for scoring. The analysis of longitudinal QoL scores will essentially be descriptive. For each of the multiple QoL scores generated from the different questionnaires, the means, medians or proportions (as appropriate) will be plotted over time together with 95% CI. These repeated measures over time may be modelled, if appropriate, with a linear or more flexible mixed model that takes account of the within-subject correlation and will allow exploration of factors associated with the outcome.

Details of the type of intervention received and complications (e.g., surgical wound infection) relating to standard of care treatments received will be monitored and recorded throughout the follow-up period and reported descriptively as frequencies and associated percentages. In relation to initial local logical diagnosis, local pathological diagnosis and integrated histological-molecular diagnosis, any difference between the tiers of diagnoses will be highlighted and categorised as: discordant; agreed; refined and reported descriptively as frequencies and associated percentages.

Confirmation of central storage of images and material, relevant targetable mutations identified by WGS and epigenomic classification, and receipt of post-mortem consent forms with confirmed central storage of samples will be recorded and reported descriptively.

Planned subgroup analyses of outcome measures include: IDH mutated and wildtype tumours; residual enhancing disease (RED; none, operable, inoperable); methylated and un-methylated tumours; age groups; types of diagnosis (WHO 2021 criteria only, epigenetic classification only, WGS analysis only, integrated diagnosis comprising all three); performance status; sex, and; biomarkers that emerge during the study (either discovered in the TJBM study or reported in the literature) that are deemed relevant after review by the Scientific Advisory Board (SAB).

### Biological samples

#### Sample collection

Biological samples will be collected at each participating site following agreed protocols and guidance from the central biospecimen coordination centre (Oxford BRAIN MATRIX Laboratory). The aim is to build as much as possible on existing GEL infrastructure and pathways. The funded pathway is represented in Figure 3.

**Figure 3:**
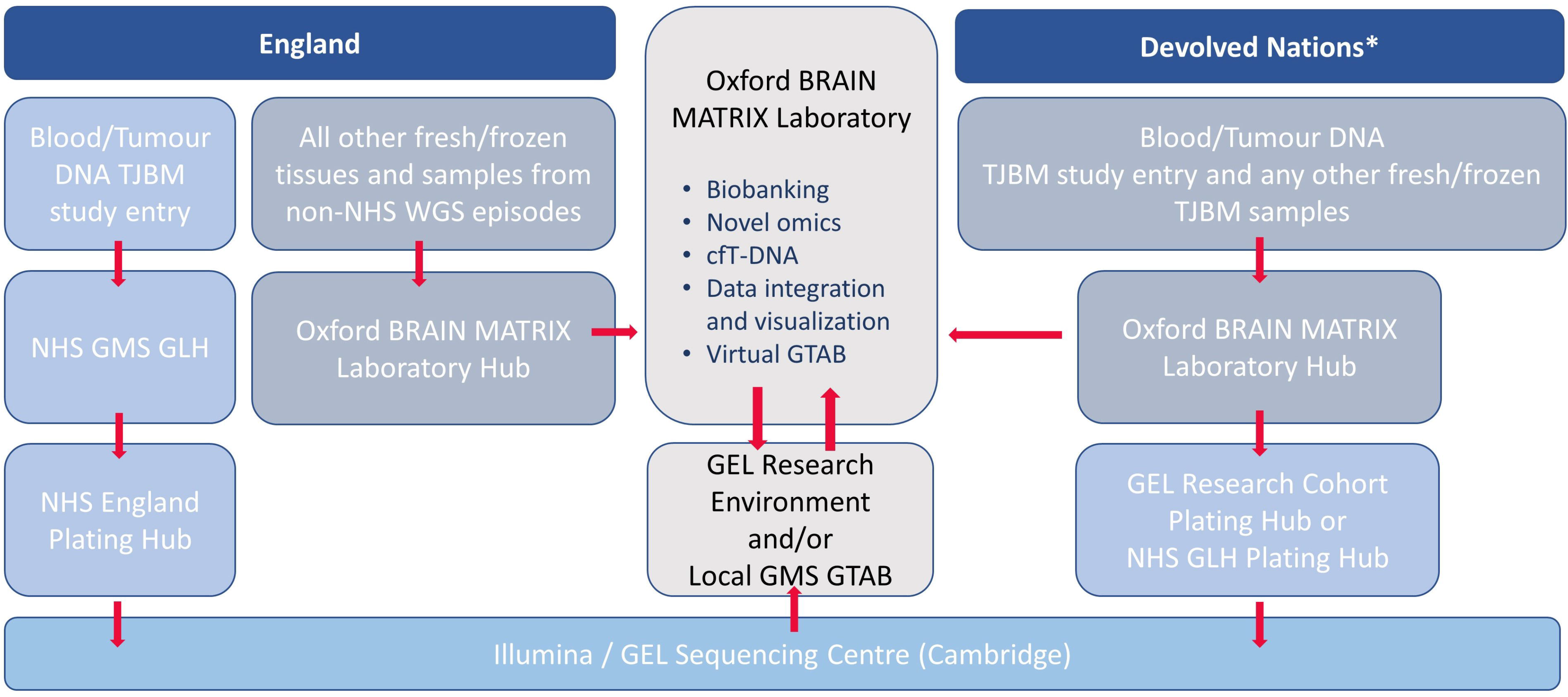
The Tessa Jowell BRAIN MATRIX Platform study sample and data flow pathways. The sample and data flow pathways within the Tessa Jowell BRAIN MATRIX Platform (TJBM) study. *TJBM sites in England are encouraged to route all samples through their local NHS GMS GLH, however, if this pathway is not yet activated or GEL consent cannot be obtained, the TJBM Study Office can facilitate the processing of samples through an alternative NHS GMS GLH or via the GEL Research pathway. GEL, Genomics England; GLH, Genomic Laboratory Hub; GMS, Genomic Medicine Service; GTAB, Genomics Tumour Advisory Board; NHS, National Health Service; WGS, Whole Genome Sequencing.

Fresh tissue for TJBM must be frozen in liquid nitrogen and stored until shipment to the Oxford BRAIN MATRIX Laboratory. Matched ‘germline’ DNA from white blood cells is required for the detection of somatic variants for paired blood/tumour WGS. This should be collected as per standard GEL protocols. Ideally, it should be collected prior to or at the time of first surgery. The blood sample(s) must be shipped together with the frozen tissue of the patient to the Oxford BRAIN MATRIX Laboratory.

For patients who are not undergoing surgery and have available tumour samples from previous tumour surgery, blood should be collected and sent to the Oxford BRAIN MATRIX Laboratory along with their tumour samples.

For participants unable to donate a blood sample, a saliva sample is an acceptable alternative.

Where possible, it is intended to collect blood samples for liquid biopsies of the tumour (e.g., tracking circulating tumour cell free (cft)DNA within the blood). Samples should ideally be collected at the time of operation, before the neurosurgeon performs any incision, ideally in theatres or the anaesthetics room just before any biopsy. Where feasible further blood samples should be collected at the following key treatment milestones:

1. At first post-operative MRI
2. Initiation of concomitant therapy (if applicable).
3. The end of concomitant therapy (if applicable).
4. Initiation of adjuvant therapy (if applicable).
5. The end of adjuvant therapy (if applicable).
6. The time of objectively measured progression.
7. During the palliative phase if a post-mortem has been agreed.

Where cerebrospinal fluid (CSF) is available for the patient, this should also be submitted.

Participating sites should follow the CRUK PEACE study protocol for post-mortem neurological tissue donation. The aim of post-mortem tissue donation is to enable research into:

1. The clonal evolution of the glioma after emergence of therapy resistance.
2. Tumour host-interaction at the whole brain level.
3. The effect of radio-chemotherapy on normal brain.
4. The interaction between the glioma and non-CNS organs (e.g., immune system in the cervical lymph nodes).

#### Sample analyses

Matched tumour/blood samples collected at first surgery are the most important samples of the TJBM study as these determine the initial integrated histological-molecular diagnosis. Following histological QC, the Oxford BRAIN MATRIX Laboratory will perform DNA extraction and QC; an aliquot will be sent to the Illumina Centre in Cambridge for WGS, and one used for EPIC methylation array at the Wellcome Trust Centre for Human Genetics in Oxford. Remaining DNA and unused tissues will be stored at the BRAIN MATRIX Biorepository.

Raw WGS data will be maintained in the GEL Data Research Environment, where it can be accessed and re-analysed by data scientists at the submitting NHS Genomic Laboratory Hub or any qualifying TJBM approved researcher. Sample data not generated by GEL will be consolidated in established bioinformatics hubs of the University of Oxford (Big Data Institute/Weatherall Institute for Molecular Medicine).

Data files on the Illumina 850k EPIC BeadChip analysis will be uploaded to the ‘Heidelberg Classifier’ hub at the German Cancer Centre in Heidelberg (27). This will generate an automated classifier report which will be stored at the Oxford BRAIN MATRIX Lab. Raw array data will be made available via the Oxford BRAIN MATRIX Laboratory.

Matched histological sections will be digitised at the Oxford BRAIN MATRIX Laboratory resulting in linked genomic data, which will initially be stored in Oxford. As the study evolves, we will aim to capture digital histological data from the material kept at the local neuropathology centre. The Oxford BRAIN MATRIX hub is working with CRUK-established data visualisation platforms, such as those developed for S:CORT (Colorectal Cancer) (28) and based on international open access platforms (such as cBioPortal (29)).

The BRAIN MATRIX neuropathology and genomics team will generate an integrated report (histology, WGS, Heidelberg Classifier) for each case in consultation with the local neuropathology team. The primary BRAIN MATRIX report will comprise the formal routine GEL WGS report (germline and tumour) and Heidelberg Classifier report and integrate this with the histological and molecular report issued by the recruiting site. It is anticipated that a local histological and molecular diagnosis using immunohistochemical surrogate markers and targeted genetic analyses will be available before WGS and Illumina EPIC BeadChip data are returned to the Oxford BRAIN MATRIX Lab. When available, the BRAIN MATRIX neuropathology and molecular genetics team will conduct a virtual MDT with the referring site to ensure all relevant information will be incorporated in the final BRAIN MATRIX diagnostic report. Variant calling and classifier outputs will be determined by the practice at the GEL/Illumina Centre and the version of the Heidelberg Classifier algorithm active at the time of sample analysis.

Where relevant germline data is identified, local sites should facilitate local genetic referral as per other GEL study protocols.

### Imaging data

Pseudo-anonymised longitudinal clinical imaging (MRI) for each patient will be collected and stored at a central imaging hub overseen by the Edinburgh Imaging Hub. Disease response assessment will be performed by practicing UK neuro-radiologists with a neuro-oncology interest via the Edinburgh Imaging Platform and additional analyses undertaken through the University of Edinburgh image analysis laboratory with permitted partners.

As per patient standard of care, it is expected that imaging will be performed pre- and post-operatively, for radiotherapy planning, following any chemoradiation, and as per follow-up determined by the managing MDT and/or if clinical concerns are raised regarding disease progression.

Pragmatic MRI protocols will be conducted as per standard of care MRI protocols and timing following diagnosis and as such will be informed by the National Institute for health and Care Excellence (NICE) guidelines from 2018 (30), in line with the recent British Society of Neuroradiologists (BSNR) imaging guidance (31), which is itself an implementation of the Brain Tumour Imaging Protocol proposed by Ellingson *et al*. (32). Where additional advanced imaging is performed this is also encouraged to be submitted to the Edinburgh Imaging Hub and will be catalogued to permit any relevant subsequent analysis. Biopsy location imaging should also be submitted, and standard operating protocols for the major neuro-navigation systems followed. Radiotherapy planning imaging will also be submitted.

The RANO assessments will be performed through a secure web portal provided by QMENTA Inc. There will be an ongoing 10% re-read to assess inter-rater agreement. It is anticipated that RANO reports will be provided within 2 weeks (10 working days) of successful transfer of imaging requiring assessment. RANO reports will categorise response where possible into Complete Response (CR), Partial Response (PR), Stable Disease (SD) or Progressive Disease (PD). In cases of diagnostic uncertainty, potential or provisional outcomes may be recorded, allowing progression events to be backdated to the correct time point should subsequent imaging be confirmatory.

### Adverse events reporting

There are no study treatments within the TJBM study. Blood sampling and completion of QoL questionnaires are the only procedures that patients undergo additional to usual care. Therefore, it is not anticipated that there will be adverse events related to participation in this study. Only severe adverse events (SAEs) relating to those additional procedures will be reported as per Common Terminology Criteria for Adverse Events (CTCAE) version 4 (33) and as defined in Supplementary Appendix 7. The reporting period is from the date of informed consent to death.

### Data management

Case report forms will be entered online via a secure web-based portal. Authorised staff at sites will require an individual secure login username and password to access this online data entry system. Paper CRFs will be available for backup only and must be completed, signed/dated and returned to the BRAIN MATRIX Study Office by the investigator or an authorised member of the site research team. Data reported on each CRF should be consistent with the source data or the discrepancies should be explained. If information is not known, this must be clearly indicated on the CRF. All missing and ambiguous data will be queried. All sections are to be completed.

All study records must be archived and securely retained for at least 25 years. No documents will be destroyed without prior approval from the sponsor, via the central Study Office. On-site monitoring will be carried out as required following a risk assessment and as documented in the Quality Management Plan. Any monitoring activities will be reported to the central BRAIN MATRIX Study Office and any issues noted will be followed up to resolution. BRAIN MATRIX will also be centrally monitored, which may trigger additional on-site monitoring. Further information regarding data management is provided in the study protocol.

The CRCTU will hold the final study dataset and will be responsible for the controlled sharing of anonymised data with the wider research community to maximise potential patient benefit while protecting the privacy and confidentiality of study participants. Data anonymised in compliance with the Information Commissioners Office requirements, using a procedure based on guidelines from the Medical Research Council (MRC) Methodology Hubs, will be available for sharing with researchers outside of the trials team within 12 months of the primary publication.

### Trial organisation structure

The University of Birmingham will act as single sponsor to this multi-centre study: Support Group, Aston Webb Building, Room 119, Birmingham, B15 2TT.). The study is being conducted under the auspices of the CRCTU, University of Birmingham according to their local procedures.

The Chief Investigator, Co-investigators, Trial Management Team Leader, Senior Trial Coordinator, Trial Coordinator, Lead and Trial Statistician, Trial Monitor and patient representatives will form the Study Management Group (SMG; current membership is listed in Supplementary Appendix 1). The SMG will be responsible for the day-to-day conduct of the TJBM study, meeting at regular intervals (e.g., at least every three months), or as required, usually by teleconference. They will be responsible for the set-up, promotion, on-going management of the study, the interpretation of the results and preparation and presentation of relevant publications.

Selected findings of clinical significance will be presented to the Scientific Advisory Board (SAB), which as a minimum will include the Chief Investigator, an oncologist, a pathologist, and a molecular biologist for a combined review of the molecular findings in context. The SAB may suggest amendments that will be incorporated into a SAB Report, which will be sent to the Executive Oversight Committee (EOC); current SAB and EOC membership is listed in Supplementary Appendix 1. Possible re-testing/further testing may be required, as a result of SAB feedback. Guided decision-making tools that review results in the context of the literature and clinical experience will be piloted within the SAB. The SAB will also evaluate research and study proposals, manage data/tissue requests, and will report back to the EOC regarding any proposals or changes which they may suggest.

The overarching remit of the EOC is to mandate, including timeframe and deliverables, the responsibility of the SAB and to take responsibility of horizon scanning to enable the incorporation of new interventional arms. They will oversee the overall study management of both the platform and any standalone interventional trials. They will also liaise with other trial units and pharma stakeholders as well as funders, charities, study sponsors, and policymakers and liaise with the BRIAN team. A quarterly EOC Report will be disseminated to all stakeholders that will demonstrate the performance metrics of each clinical site. In addition, the use of all samples given to external researchers via the SAB will be included in the report.

### Confidentiality

Confidential information collected during the study will be stored in accordance with the General Data Protection Regulation (GDPR) 2018. As specified in the patient information sheet (PIS) and with the patients’ consent, patients will be identified using only their date of birth and unique study ID number. Authorised staff may have access to the records for quality assurance and audit purposes. The BRAIN MATRIX Study Office maintains the confidentiality of all patients’ data and will not disclose information by which patients may be identified to any third party other than those directly involved in the treatment of the patient and organisations for which the patient has given explicit consent for data transfer (e.g., laboratory staff).

### Trial status

Recruitment for the study opened in Nov-2020 and recruitment is expected to last for 5 years.

## Ethics and dissemination

The study will be performed in accordance with the recommendations guiding physicians in biomedical research involving human subjects, adopted by the 18^th^ World Medical Association General Assembly, Helsinki, Finland and stated in the respective participating countries laws governing human research, and Good Clinical Practice. The protocol was initially approved on 18-Feb-2020 by West Midlands - Edgbaston Research Ethics Committee; the current protocol (v3.0) was approved on 15-Jun-2022.

A meeting will be held after the end of the study to allow discussion of the main results among the collaborators prior to publication. The results of this study will be disseminated through national and international presentations and peer-reviewed publications.

Manuscripts will be prepared by the SMG and authorship will be determined by mutual agreement.

## Discussion

### Justification for patient population

The main aim of the TJBM study is to test the hypothesis that comprehensive genomic and epigenomic profiling of gliomas is feasible in a timely manner in the UK, and that the results improve stratification of patients for next generation (targeted) therapies, ultimately improving outcomes and QoL.

Gliomas occur at all ages and their specific subtype is difficult to predict preoperatively. Therefore, the patient population eligible for the TJBM study is broad. It includes any patient who on preoperative assessment is suspected to have a diffuse glioma, or where a diffuse glioma remains a credible differential diagnosis, as established by the multidisciplinary team at the recruiting site.

Recruitment is also open to patients with diffuse or atypical gliomas who had a biopsy before the launch of the TJBM study. This approach ensures that:

1. Patients with slow-growing diffuse gliomas that evolve over many years (oligodendrogliomas) may benefit from one or more of the clinical trials developed as part of the TJBM study and sample collection at tumour progression.
2. Patients who potentially benefit most from comprehensive molecular diagnostics are not missed, which often are those with rare or atypical variants of glioma based on current diagnostics (34).

The infrastructure for children with brain tumours in the UK is different to that of adults with epigenomic classification regularly used and with WGS shortly to be made available for all children with cancer. Thus, the challenges and opportunities are different for paediatric patients. Nevertheless, it is intended that children will be included within the TJBM study wherever appropriate. The recruitment of children and adolescents into the TJBM study will be coordinated with the UK’s paediatric oncology expert group(s).

### Justification for methodology

Neuroimaging with MRI plays a central role in the initial diagnosis and treatment stratification, surgical and radiotherapy planning and assessment of disease response and progression in glioma.

Potential participants will have undergone MRI as part of a diagnostic work-up and an intracranial mass identified. As per current clinical practice, expert neuroradiological review will indicate glioma as the likely diagnosis, and a plan for treatment will be made by the relevant regional MDT meeting. Those with suspected glioma on MRI who are to undergo surgery and are otherwise eligible for the TJBM study will be approached. A proportion of these lesions that are subsequently diagnosed as types of brain tumours other than glioma will inevitably be included. They will not be included in the main analysis; however, data from these will be stored for additional research into those less common tumours which can be challenging to diagnose from imaging alone.

At the time of surgery, neuro-navigation will be used to capture the location of each biopsy taken for diagnosis and further molecular characterisation within the TJBM study. This will aid with subsequent analysis of tumour molecular markers and heterogeneity in the context of radio-genomics – a recent development in cancer imaging for assessment of disease in the personalised medicine era employing machine learning and artificial intelligence approaches (32).

Early post-operative imaging for suspected HGG, including pre- and post-contrast T1-weighted MRI, is currently recommended within 72 hours of surgery to minimise the presence of non-neoplastic enhancement. This imaging will be captured to permit eventual assessment of extent of resection, or the volume of residual enhancing disease, important prognostic factors for glioma. Subsequent to this, any additional imaging for radiotherapy planning, including computerised tomography (CT), will be captured. Under the most recent response assessment in neuro-oncology (34) recommendations for HGG from 2017, this is recommended as the baseline for assessing treatment response (32).

All imaging will receive primary radiological reports at the local site as per current standard clinical practice. To deliver the imaging outcome requirements for clinical trials, centralised RANO reads will be delivered by a core group of UK Consultant neuro-radiologists with a subspecialty interest in neuro-oncology imaging from TJBM study sites. Radiotherapy planning data in the form of dose distribution maps and associated data will be integrated into the imaging database to permit subsequent analysis of spatiotemporal response patterns of different glioma-treatment combinations in light of received radiation dose.

Local radiological reporting will inform clinical decision making for individual patients. Central image analysis will provide resilient standardised assessment to meet internationally-accepted standards which are suitable for peer-reviewed publication and are accepted by major regulatory approval bodies. All relevant pre-operative and subsequent imaging will be identified for pseudonymisation and uploaded to the BRAIN MATRIX Imaging Platform in Edinburgh. This will be achieved through a dedicated online secure portal, with alternative secure online and physical transmission pathways available for redundancy. The accrued imaging data will form a core resource that will be leveraged for future imaging and clinical radiological research. The platform can also provide the basis for additional imaging studies within non-standard of care imaging and advanced techniques, which would be separately funded and detailed in their respective project documentation.

### Justification for tissue collection

Historical approaches to brain tumour tissue collection in formaldehyde and paraffin are not currently fully compatible with modern genomic technologies, which require frozen tissue. This is why the collection of fresh frozen (FF) material is essential for the TJBM study. Pairing with non-neoplastic, so-called germline DNA, is also essential for confident calling of somatic variants in the tumour. Germline DNA analysis will also provide novel data on genetic risk for glioma predisposition.

Blood plasma, where possible, will be collected to facilitate future analysis of cftDNA. This technology is still in its infancy; however, it is clear that non-invasive, real-time monitoring of tumour evolution will become feasible in the next 5-10 years (35). Similarly, where available for the patient, CSF will be submitted.

Post-mortem tissue banking via the CRUK PEACE and MRC Brain BioLink projects will allow us to study the glioma-brain interface, extent of spatial tumour heterogeneity, genomic signature of the treatment resistant tumour clones, and effect of treatments on normal brain. Systematic post-mortem brain banking for research into adult gliomas does not currently exist in the UK.

### Justification for molecular diagnostics

All TJBM study baseline diagnoses will follow the new WHO classification of tumours of the CNS from 2021 to achieve an integrated histological-molecular diagnosis for diffuse gliomas (10). This is achieved with a combination of immunohistochemical surrogate markers and – in most instances – targeted or panel sequencing for relevant hotspot mutations, or cytogenetics (7). Only in exceptional circumstances unbiased ‘omics’ approaches are used, such as the epigenomic ‘Heidelberg Classifier’ (34). Further, any diagnostic approach may differ between centres in the UK, making analyses of cohorts pooled from different sites difficult.

The new WHO 2021 classification represents a consensus opinion based on new insights into the molecular definition of brain tumours from cIMPACT-NOW. cIMPACT-NOW updates are not intended to supplant the existing WHO classification, but to provide possible guidelines for practicing diagnosticians and future WHO classification updates. It is clear from the first iterations of cIMPACT-NOW that any progress is driven by next generation (‘omics’) molecular analysis, not by standard or targeted analyses. This insight underpins the selection of molecular analytical tools for the TJBM platform, namely, combined paired (blood-tumour) WGS DNA analysis integrated with the epigenomic ‘Heidelberg Classifier’ (34). To achieve this, the TJBM study will build on the experience and UK infrastructure of the 100,000 genomes project led by GEL, which introduced WGS pathways into clinical practice. Early results from WGS in non-brain cancer patients suggest that virtually all patients could be mapped to existing or potential targeted therapies (36). Prospective large-scale paired WGS sequencing studies in diffuse glioma patients have not been done; however, experience from other brain tumours such as medulloblastoma, suggest that analysis of WGS data will provide new insights into glioma subtype diversity, including alterations in specific non-coding regulatory elements not evident from non-WGS genomic approaches (37). WGS data will capture all currently known relevant variants and uncover novel variants relevant for a better understanding of tumour evolution and response to treatment. WGS and epigenomic analyses are highly complementary genomic approaches (37): WGS will establish all potentially actionable mutations and epigenomic classification will establish an unbiased score for the precise classification of the glioma (34). Neither can be achieved with conventional targeted sequencing approaches. Importantly, the epigenomic classification by novel DNA methylation-based ‘Heidelberg Classifier’ has been shown to fundamentally alter clinical diagnoses and histological grades in >10% of biopsies, leading to changes in therapy (34). Moreover, the epigenomic array technology provides genomic copy number variant data, which in theory can also be inferred from WGS data. However, pipelines for this type of analysis from WGS data are just evolving and it is predicted that comparative analysis of WGS and Illumina’s EPIC BeadChip data will be bioinformatically highly valuable. Finally, WGS and the EPIC array raw data will form a unique source for researchers who will be able to access this data together with all clinical, imaging and histological data. Creating a relatively future-proof, quality-controlled research infrastructure is one of the main aims of the TJBM study, in addition to establishing feasibility of timely genomic diagnosis in the NHS setting.

the TJBM study is more than paired tumour/blood WGS and EPIC array analysis. As the former is being rolled out in the NHS in England, the BRAIN MATRIX molecular neuropathology team will work with GEL and other stakeholders (including industry) to explore the next-generation of tissue analytics (such as long-read sequencing and mass spectrometry). This will result in a unique, prospectively acquired dataset that will enable researchers to integrate data analysis across modalities and with outcomes and treatment response.

### Justification for patient outcome and quality of life

Clinical trials require prospective collection of clinical data to Good Clinical Practice (GCP) standards. This platform will develop the infrastructure for the collection of this, enabling streamlined patient recruitment into clinical trials. Evaluation of treatments and associated complications across patients will give an accurate measure of adverse events associated with current standard of care treatment in practice in the UK.

Maximising QoL for patients with diffuse glioma is important particularly given its poor prognosis; therefore, standardised measures of QoL will be collected. In adults these have been selected for their standardisation and ease of use and have been reviewed with input from Patient Public Involvement (PPI) representatives. It is intended that in the future integration with The Brain Tumour Charity’s BRIAN project (REC Reference: 18/SC/0283) (19) will allow further understanding of patient reported outcome measures.

## Data Availability

All data produced in the present study are available upon reasonable request to the authors

## Contributors

CW (Chief Investigator and first author): conception and design of the study, drafting and review of paper.

JS: trial management team leader, study design, drafting and review of paper.

AP: Study Biostatistician, study design, drafting and review of paper.

RM: senior trial coordinator, review of paper.

VW: principal investigator (Birmingham), review of paper.

UP: neuropathologist (Birmingham), study design, review of paper.

HB: patient and public representative, study design, review of paper.

JA: study design, review of paper.

RS: design of the study, review of paper.

GT: conception and design of the study, review of paper.

AM: conception and design of the study, review of paper.

OA: conception and design of the study, review of paper.

LB: Lead Biostatistician, design of study, drafting and review of paper.

## Acknowledgements

We would like to thank our PPI representative, Peter Buckle who, with Helen Bulbeck, has been instrumental in making sure the patients’ voice has been heard and incorporated into this study protocol. In addition, we thank staff past and present from the CRCTU, University of Birmingham including Clive Stubbs, Richard Fox, Dr Sarah Bowden, Prof. Christina Yap, Dr Siân Lax and Dr Louisa Jeffery, and Hannah Brooks at the University of Oxford for contributions to the protocol and paper.

## Additional Files

**Supplementary Appendix 1: Tessa Jowell BRAIN MATRIX Platform Study Investigators & Committee Membership**

Principal Investigators, Study Management Group (SMG), Executive Oversight Committee (EOC) and Scientific Advisory Board (SAB).

**Supplementary Appendix 2: The Tessa Jowell BRAIN MATRIX Platform study SPIRIT Checklist**

A completed Standard Protocol Items: Recommendations for Intervention Trials (SPIRIT) checklist for the Tessa Jowell BRAIN MATRIX Platform study protocol.

**Supplementary Appendix 3: The Tessa Jowell BRAIN MATRIX Platform study World Health Organization Trial Registration Data Set**

The World Health Organization (WHO) trial registration data set for the Tessa Jowell BRAIN MATRIX Platform study.

**Supplementary Appendix 4: The Tessa Jowell BRAIN MATRIX Platform study informed consent form**

The informed consent form for the Tessa Jowell BRAIN MATRIX Platform study.

**Supplementary Appendix 5: The Tessa Jowell BRAIN MATRIX Platform study lay summary and patient information sheet**

The lay summary and patient information sheet for the Tessa Jowell BRAIN MATRIX Platform study.

**Supplementary Appendix 6: The Tessa Jowell BRAIN MATRIX Platform study schedule of events**

Patient schedule of events for the Tessa Jowell BRAIN MATRIX Platform study.

**Supplementary Appendix 7: Definition of adverse events**

Definitions of adverse events used for the Tessa Jowell BRAIN MATRIX Platform study.

