## Supplementary Appendix 1 for "Protocol for the Tessa Jowell BRAIN MATRIX Platform Study"

### Supplementary Appendix 1: Tessa Jowell BRAIN MATRIX Platform Study Investigators & Committee Membership

**Principal Investigators**

- Dr Victoria Wykes - Queen Elizabeth Hospital, Birmingham
- Prof. Keyoumars Ashkan - King's College Hospital, London
- Prof. Michael Jenkinson - The Walton Centre, Liverpool
- Prof. Susan Short - St James's University Hospital. Leeds
- Dr Paul Brennan - NHS Lothian
- Dr Richard Mair - Addenbrooke's Hospital, Cambridge
- Dr Stuart Smith - Queens Medical Centre, Nottingham
- Prof. Anthony Chalmers - Greater Glasgow and Clyde
- Dr David Coope - Salford Royal Hospital, Manchester
- Dr Catherine McBain - The Christie, Manchester
- Dr Clare Hobbs - John Radcliffe Hospital, Oxford

**Study Management Group (SMG)**

- Professor Colin Watts (Chief Investigator) - Institute of Cancer and Genomic Sciences (ICGS), University of Birmingham
- Professor Olaf Ansorge - University of Oxford
- Dr Ute Pohl - University Hospitals Birmingham
- Professor Adam Waldman - University of Edinburgh
- Dr Gerard Thompson - University of Edinburgh
- Dr John Apps - University of Birmingham
- Dr Victoria Wykes - University of Birmingham
- Professor Lucinda Billingham – CRCTU, University of Birmingham
- Dr Rowena Sharpe – CRCTU, University of Birmingham
- Amit Patel - CRCTU, University of Birmingham
- Dr Joshua Savage - CRCTU, University of Birmingham
- Rhys Mant - CRCTU, University of Birmingham
- Dr Louisa Jeffery - CRCTU, University of Birmingham
- Hannah Brooks – University of Oxford
- Peter Buckle (PPI Representative)
- Dr Helen Bulbeck (PPI Representative) - brainstrust

**Executive Oversight Committee (EOC)**

- Professor Tim Maughan (Chair)– Professor of Clinical Oncology, CRUK/MRC Institute for Radiation Oncology, University of Oxford

- Professor David Cameron – Clinical Director of the Cancer Research UK Edinburgh Centre, University of Edinburgh
- Dr Helen Campbell – Portfolio Manager for Department of Health and Social Care Research Networks, Clinical Research Facilities, and Cancer Research, University of Exeter
- Dr Mark Gilbert – Chair of the Neuro-Oncology Branch, Centre for Cancer Research, National Cancer Institute
- Professor Richard Gilbertson – Director of CRUK Cambridge Centre
- Professor Max Parmar – Director of the MRC Clinical Trials Unit, UCL
- Lord James O'Shaughnessy – Member of House of Lords
- Professor Steven Pollard – Group Leader, MRC Centre for Regenerative Medicine and Edinburgh Cancer Research Centre, University of Edinburgh
- Jess Mills – Co-Founder and Special Advisor, Tessa Jowell Brain Cancer Mission

**Scientific Advisory Board (SAB)**

- Professor Ruth Plummer (Chair) - Professor of Experimental Cancer Medicine, Newcastle University
- Professor Neil Carragher - Professor of Drug Discovery, University of Edinburgh
- Professor Anthony Chalmers - Chair of Clinical Oncology, University of Glasgow
- Will Jones - Chief Executive, brainstrust
- Dr Juanita Lopez - Consultant Medical Oncologist, The Royal Marsden NHSFT
- Professor Whitney Pope – Director, Brain Tumour Imaging, UCLA
- Professor Thomas Wurdinger - Professor of Neurosurgery, Amsterdam UMC
- Professor Christina Yap - Professor of Clinical Trials Biostatistics, ICR
- Professor Patrick Wen - Professor of Neurology, Harvard Medical School
- Professor Roel Verhaak - Professor and Associate Director for Computational Biology, The Jackson Laboratory for Genomic Medicine
