## Supplementary Appendix 3 for "Protocol for the Tessa Jowell BRAIN MATRIX Platform Study"

### Supplementary Appendix 3: The Tessa Jowell BRAIN MATRIX Platform study World Health Organization Trial Registration Data Set

| **Data category** | **Information** |
|---|---|
| Primary registry and trial identifying number | ClinicalTrials.gov<br>NCT04274283 |
| Date of registration in primary registry | 18-Feb-2020 |
| Secondary identifying numbers | ISRCTN 14218060 |
| Source(s) of monetary or material support | The Brain Tumour Charity<br>Genomics England<br>INNOVATE UK |
| Primary sponsor | University of Birmingham |
| Secondary sponsor(s) | n/a |
| Contact for public queries | Dr Joshua Savage: |
| Contact for scientific queries | Prof. Colin Watts: |
| Public title | The Tessa Jowell BRAIN MATRIX Platform Study |
| Scientific title | A British Feasibility Study of Molecular Stratification and Targeted Therapy to Optimize the Clinical Management of Patients With Glioma by Enhancing Clinical Outcomes, Reducing Avoidable Toxicity, Improving |

| Data category | Information |
|---|---|
| | Management of Post-operative Residual & Recurrent Disease and Improving Survivorship |
| Countries of recruitment | UK |
| Health condition(s) or problem(s) studied | Newly diagnosed suspected, or progressive with known, WHO Grade 2-4 glioma |
| Intervention(s) | None |
| Key inclusion and exclusion criteria: | Ages eligible for study: ≥16 years<br>Sexes eligible for study: both<br>Accepts healthy volunteers: no |
| | Inclusion criteria: Newly diagnosed suspected, or progressive with known, WHO Grade 2-4 glioma |
| | Exclusion criteria: Primary spinal cord tumours, active treatment of other malignancy |
| Study type | Observational |
| | Primary purpose: Feasibility of establishing an integrated histological-molecular diagnosis of glioblastoma |
| Date of first enrolment | 14-Dec-2020 |
| Target sample size | 1000 |
| Recruitment status | Open |
| Primary outcome(s) | • Time (from biopsy) to integrated histological–molecular diagnosis (TTMD) using whole genome sequencing and epigenomic classification |

| **Data category** | **Information** |
|---|---|
| Key secondary outcome(s) | • Time to completion of each node of tissue and imaging pathway.<br>• Tumour and biological sample(s) quality control (QC) status.<br>• Imaging QC status.<br>• Inter-rater agreement of RANO assessments. |
