## Supplementary Appendix 4 for "Protocol for the Tessa Jowell BRAIN MATRIX Platform Study"

*To be printed on hospital headed paper*

### TESSA JOWELL BRAIN MATRIX

**A BRitish feasibility study of molecular stratification and targeted therapy to optimize the clinical mAnagement of patIeNts with glioMA by enhancing clinical ouTcomes, Reducing avoIdable toXicity, improving management of post-operative residual & recurrent disease and improving survivorship**

#### Platform Study

### Informed Consent Form

| | | | |
|---|---|---|---|
| **Site:** | ______________________ | **Patient's Study Number:** | ☐☐☐☐ |
| **Principal Investigator:** | ______________________ | | |

**Sponsor's Protocol Number: RG_18-258**

**Please initial each box**

| | | |
|---|---|---|
| **1.** | I confirm that I have read and understand the Patient Information Sheet (version ................... dated..........................) for the above study. I have had the opportunity to consider the information, ask questions and have had these answered satisfactorily. | ☐ |
| **2.** | I understand that my participation is voluntary and that I am free to withdraw at any time without giving any reason, without my medical care or legal rights being affected. | ☐ |

3. I give my permission for my initials, date of birth, hospital number, NHS/CHI number, Genomics England Referral ID number, histopathology numbers, and radiology numbers to be given to the BRAIN MATRIX Study Office when I am registered to the study as well as a copy of this consent form. I understand that copies of the consent form may be forwarded to other healthcare professionals to prove that I am taking part in the study. I also give permission for my information to be shared with other hospitals involved in my care and with the National Cancer Registration Service.

4. I understand that relevant sections of my medical notes and data collected during the study may be looked at by individuals from the BRAIN MATRIX Study Office, regulatory authorities, Sponsors and/or NHS bodies, where it is relevant to my taking part in this research. I understand that this information will be held in a confidential manner. I give permission for these individuals to have access to my records.

5. I agree to the collection of blood samples, or any other biofluid samples, from me, and consent for these samples to be analysed (including genetic analysis) as part of research associated with this study, to be used in biobanking and for future ethically approved studies.

6. I agree to the collection of tissue from any of my previous biopsies and future tumour surgeries (including "surgical access tissue") and consent for these samples to be analysed (including genetic analysis) as part of research associated with this study, to be used in biobanking and for future ethically approved studies.

7. I understand that identifiable data from the study may be shared with other institutions involved in BRAIN MATRIX (University of Edinburgh, University of Oxford, Genomics England, BRIAN databank run by The Brain Tumour Charity).

8. Anonymised linked data may also be provided to other 3rd parties (e.g. other academic institutions, pharmaceutical companies and QMENTA) for research and safety monitoring purposes and may be placed in recognised medical research data repositories, in accordance with best research practice.

9. I agree to my GP being informed of my participation in this study and to my GP being sent a copy of this Patient Information Sheet.

10. I agree to the collection of different aspects of my health data from the NHS and other Department of Health organisations, in addition to my medical records. The collection and analysis of my health data for research will continue across my entire lifetime and beyond.

11. I agree to take part in the above study.

**The following are optional and will not affect entry into the study, please initial in the relevant box:**

| | No | Yes |
|---|---|---|
| If in the event that an abnormality that might affect other family members is uncovered during genetic testing for the study, I would like my doctor to be informed and to be referred to a genetic counsellor if appropriate. | | |
| I would like to receive a copy of the lay summary results at the end of the study. | | |
| I agree that my doctor, in consultation with the BRAIN MATRIX Study Office, together with Genomics England and other NHS staff, can contact me if the data or samples reveal any clinical trials or other research that I might benefit from. If something is relevant to me, there is a process by which this will be shared with my NHS clinical team. | | |

| | | |
|---|---|---|
| **Name of patient** | **Date** | **Signature** |
| **Name of person taking consent**<br>You must have signed the<br>Site Signature & Delegation Log | **Date** | **Signature** |
