## Supplementary Appendix 5 for "Protocol for the Tessa Jowell BRAIN MATRIX Platform Study"

**A BRitish feasibility study of molecular stratification and targeted therapy to optimize the clinical mAnagement of patIeNts with glioMA by enhancing clinical ouTcomes, Reducing avoIdable toXicity, improving management of post-operative residual & recurrent disease and improving survivorship**

#### Platform Study

The **Tessa Jowell BRAIN MATRIX** is a programme of work, the principal purpose of which is to improve the knowledge of, and treatment for, glioma, a type of brain tumour.

Brain tumours arise due to changes in the DNA and other molecules in cells of the brain. Different types of gliomas can have different changes and these can be used to determine a precise ‘molecular diagnosis’. The ultimate goal for the Tessa Jowell BRAIN MATRIX is to learn how to use these molecular changes to more precisely determine what exact type of tumour patients have, and to identify, decide and test whether specific ‘targeted’ treatments could improve the survival and/or quality of life of patients with brain tumours.

The Tessa Jowell BRAIN MATRIX Platform Study forms the backbone of this programme. You have been given this information because you either have an existing brain tumour, or have recently been diagnosed with a brain tumour, likely to be a glioma.

In the platform study we aim to develop the infrastructure to provide rapid and accurate molecular diagnosis and the infrastructure to deliver clinical trials of new therapies in the future.

**The diagram below shows a summary of the Tessa Jowell BRAIN MATRIX Programme:**

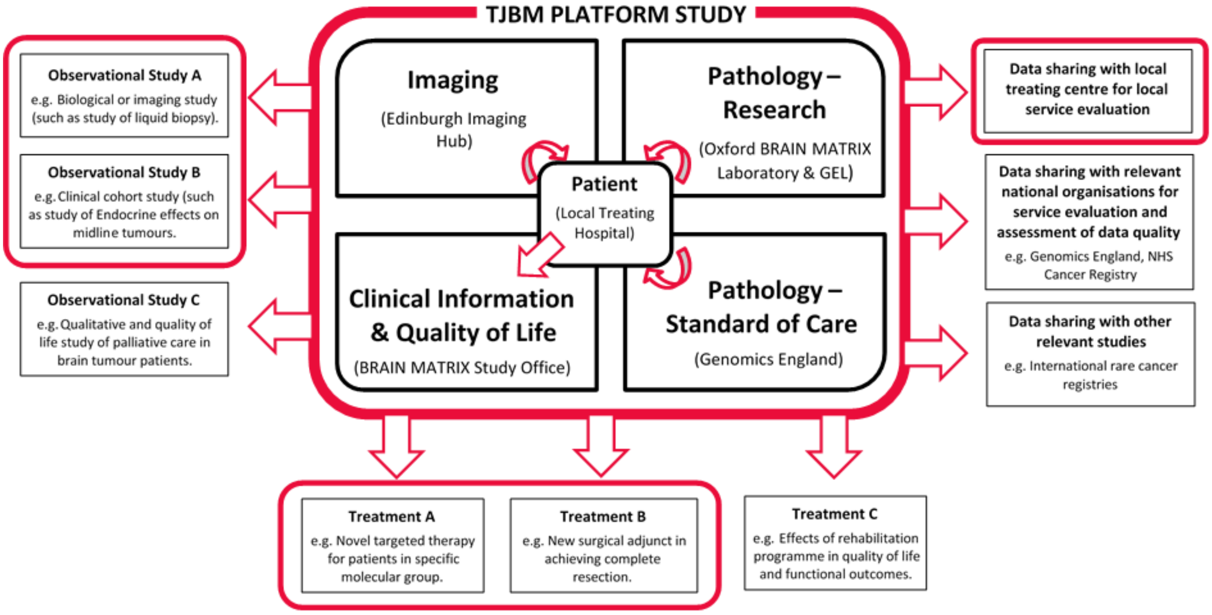


For further information about the Tessa Jowell BRAIN MATRIX Platform Study, please ask your doctor.

*To be printed on local hospital headed paper*

### TESSA JOWELL BRAIN MATRIX

**A BRitish feasibility study of molecular stratification and targeted therapy to optimize the clinical mAnagement of patIeNts with glioMA by enhancing clinical ouTcomes, Reducing avoIdable toXicity, improving management of post-operative residual & recurrent disease and improving survivorship**

#### Platform Study

### Patient Information Sheet

*To be used for patients over 16 years of age considering entering the Platform Study*

#### We invite you to take part in a clinical study

We would like to invite you to take part in a non-commercial clinical study run by the University of Birmingham, supported by The Brain Tumour Charity.

Before you decide whether you would like to participate in this study, we would like you to understand:

- Why the research is being done.
- What it would involve for you.

A member of the team at your local hospital involved in this study will go through this information sheet with you and give you a copy to take away with you to discuss with friends and/or relatives, if you wish. They will answer any questions that you may have. If there is anything that is not clear within the information sheet, please ask your medical team.

**Take your time to decide whether or not you wish to take part. If you decide not to take part your doctors will continue to treat you in line with standard treatment and it will not affect the quality of your care.**

#### Contents


**Part 1: What is this clinical study about?**

1. What is the purpose of this study?
2. What will happen to me if I take part?
3. What are the possible benefits and risks of taking part?
4. What happens at the end of the study?
5. Will anybody get paid if I take part?
6. What if there is a problem?
7. Will my taking part in this study be kept confidential?

**Part 2: What else do I need to know about taking part in this clinical study?**

1. More information about taking part
2. Who should I contact for further information

#### Important things that you need to know

We want to improve the outcome for people like yourself who are living with a brain tumour. Specifically, in this study we are establishing a platform to deliver:

- Rapid and accurate detailed laboratory analysis of the molecular changes in brain tumours. This will enable accurate diagnosis in a timeframe useful to clinicians.
- To develop robust ways of collecting, processing and storing brain tumour tissue, radiology images and clinical data.
- An enhanced network of UK brain tumour clinical centres.
- The introduction of clinical trials of therapies tailored/targeted to the precise molecular and clinical features of individual patients.

#### Who is Tessa Jowell?

This study is named after Baroness Tessa Jowell, the ex-politician and minister, who died from a glioma in summer 2018. As a result of her efforts to raise awareness of brain tumours, research funding is now being increased.

#### Abbreviation and definitions

- Biofluids — Bodily samples that are liquid (e.g. blood, urine, saliva, tears, or cerebrospinal fluid
- DNA — Deoxyribonucleic acid
- GDPR — General Data Protection Regulation
- GEL — Genomics England
- Glioma — A type of brain tumour
- NHS — National Health Service
- REC — Research Ethics Committee
- TJBM — Tessa Jowell BRAIN MATRIX

We have sought to make this document readable and understandable by patients by involving patient representatives in writing this document. However, due to the nature of the study, and the legal and regulatory requirements to include certain information, we are aware that some of the wording can seem complex. This makes it especially important that you ask your research team about anything that you do not fully understand.

If you have any further questions, you are very welcome to contact the research team.

### Part 1:
### What is this clinical study about?

#### 1. What is the purpose of this study?

##### What is the purpose of this study?

The **Tessa Jowell BRAIN MATRIX** is a programme of work, the principal purpose of which is to improve the knowledge of, and treatment for, glioma, a type of brain tumour. The platform study forms the backbone for this.

Brain tumours are a challenging disease to treat. This is due to:

- The tumour's location within the brain, and its tendency to grow into nearby brain tissue, often make it very difficult or not possible to remove the tumour completely with surgery.
- The biology of brain tumours, including:
  - Their lack of response to traditional radiotherapy/chemotherapies.
  - Their ability to spread within the brain.
  - Their ability to change/adapt during/in response to treatments.
- Difficulties in delivering drugs in adequate amounts to the tumour, due to natural defences of the brain (the 'blood-brain barrier').
- A relative lack in research infrastructure for brain tumour patients within the NHS and academia in the UK.

Together this means that progress in treating brain tumours has not matched that of other cancers. In this study, we want to create an underlying treatment and analysis 'platform' to address these challenges of treating gliomas. This platform will bring together many different people including patients like you, brain surgeons, cancer doctors, technicians and scientists.

The standard treatment for most brain tumours is surgery. This is to remove the main bulk of the tumour and is followed by radiotherapy and/or chemotherapy. In the majority of cases, however, it is difficult for all of the tumour cells to be removed by the surgeon because of the way it invades, and the potential damage that can occur to the brain by removing too much tissue. Therefore, some tumour cells can be left behind which can regrow. Whilst radiotherapy and/or chemotherapy can delay this, their effects are often only temporary. There is, therefore, a need to develop and test new treatments.

Brain tumours arise due to changes in the DNA and other molecules in cells of the brain. Different types of gliomas can have different changes and these can be used to determine a precise 'molecular diagnosis'. The ultimate goal for the **Tessa Jowell BRAIN MATRIX** is to learn how to use these molecular changes to more precisely determine what exact type of tumour you have and to identify, decide and test whether specific 'targeted' treatments could improve the survival and/or quality of life of patients with brain tumours.

To achieve this, we need to develop the infrastructure to provide a rapid and accurate molecular diagnosis. A large network of clinical hubs across the UK, with expertise in managing patients with brain tumours, will be developed. This will enable:

- Collection of high-quality clinical data, including information about your quality of life, in a secure manner, maintaining patient confidentiality.
- Robust processes and safeguards for the collection, processing, analysis and storage of tumour tissue, including feedback to your treating doctors of a precise molecular diagnosis.
- Collection and expert review of images to assess responses of tumours to treatment in a standardised manner and in a time frame relevant to a patient's treatment.

Once established this infrastructure will facilitate the rapid introduction of clinical trials testing targeted therapies tailored to the genetic changes of an individual's tumour. In addition, collection of this data, images and samples, will provide an extremely valuable resource to help us to understand better the clinical behaviour, impact on patients, molecular pathology and imaging characteristics of brain tumours. This will enable clinicians and scientists to improve the quality of treatments available and the quality of lives of patients with brain tumours. This study will actively engage with partners within the NHS, universities, pharmaceutical industry and charitable sectors to translate the advances into benefits for patients.

##### Why have I been invited to participate?

In most cases, you have been invited to take part in this study because your recent scan shows that there is a growth in your brain. It is likely that this is a glioma, a type of brain tumour, and that you will be undergoing surgery (or biopsy) to remove at least some of this tumour.

In some cases, you have been asked because you are known to have such a tumour, and are undergoing further surgery, or samples are available that can be used from your previous operations.

##### Do I have to take part?

No, you do not have to take part. It is up to you to decide whether to join the study. A member of the local research team will describe the study and go through this information sheet with you. If you decide to take part, you will be asked to sign a consent form to show you have agreed to participate. You are free to withdraw at any time, without giving a reason. A decision not to take part or to withdraw later will not affect the standard of care you receive thereafter.

#### 2. What will happen to me if I take part?

##### What will happen if I decide to take part?

If you decide to take part in this study, you will be given this information sheet to keep, and you (and your study doctor) will be asked to sign a consent form to show that you have agreed to take part in this study. You will be given a copy of the signed consent form to take home with you. We will then need to confirm that you meet all the requirements and that you are fit enough to take part in the study. This is called screening. If you do not meet the inclusion criteria for this study you will not be able to participate. Decisions about your treatment are independent to your participation in the BRAIN MATRIX Platform Study and will continue to be made in discussion with you and your doctor.

**The diagram below shows a summary of the Tessa Jowell BRAIN MATRIX Programme:**

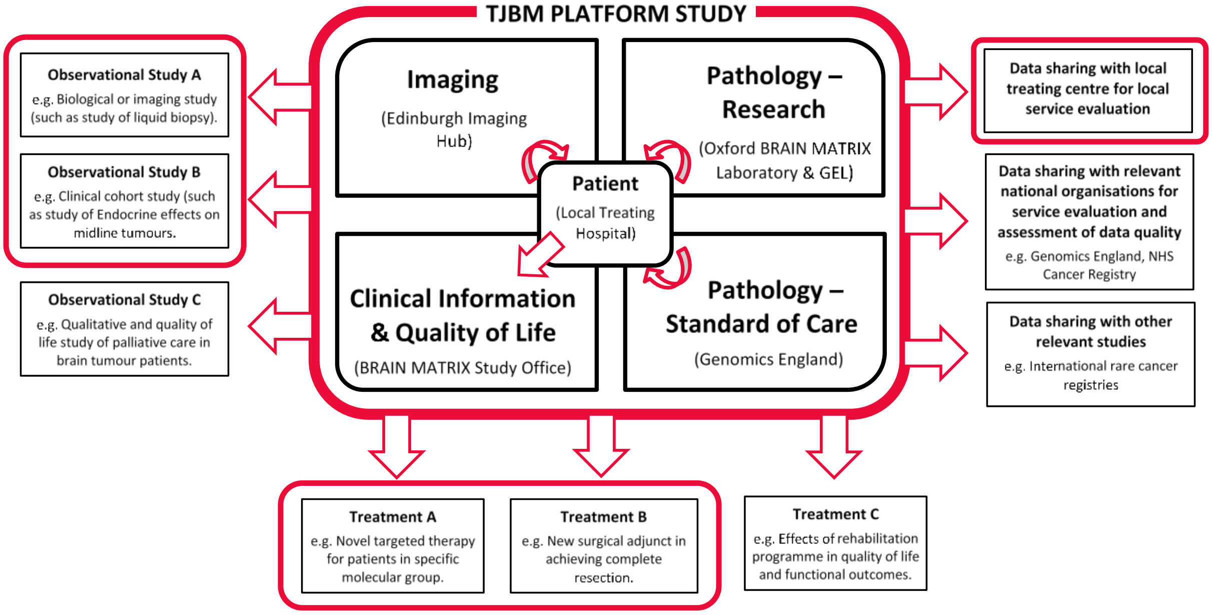


#### Study Participation

If you are eligible for this study you will either have had, or be about to have, surgery for your tumour. As part of this study, tumour removed during your operation will be analysed to look for specific molecular changes. No additional surgery or extra tumour will be taken as a result of participation in this study. As with normal standard care, your tumour will be analysed by your local pathologist. A small part will be sent for review by experts and advanced molecular analysis will be undertaken (in most cases analyses called whole genome sequencing and epigenetic classification) to get a detailed understanding of the DNA/molecular changes within your tumour. These results will be fed back to the doctor treating you. It is intended that this will occur within 28 days; however, it may be longer while the study becomes fully operational. If samples are available from previous surgery to your tumours, we may also analyse these. Similarly, if available, other relevant samples, such as cerebrospinal fluid, collected as part of your care, may also be analysed. In addition, as technologies and analyses improve our understanding of brain tumours, we may find important results at a later date. These will be fed back to your doctor.

To perform these analyses you will be asked to give a blood sample, which will also be analysed to look at the molecular features, including of your DNA. This is required to identify what 'new' changes have occurred in your tumour.

Following your surgery you will continue with other treatment(s) as directed by your doctor. Treatment generally involves radiotherapy and chemotherapy and your doctor will discuss this with you. As is standard practice, you will be closely monitored for signs of disease progression and the effects of the treatment given.

As part of this study, information on your treatments and disease will be collected. This will include details about, your current health (e.g. as assessed by your doctor's assessment and examination, some blood test results), treatments you are taking/receiving (e.g. names of drugs, doses). Images from brain scans you undergo, along with relevant clinical information, will also be sent to and stored by the University of Edinburgh, and where appropriate, undergo expert review by a panel of radiologists with expertise in brain tumours. You will not have any additional scans as part of this study. If you have further surgery, some of the tissue removed may also be analysed. As above, no additional surgery will be performed or extra-tumour will be removed as a result of participation in this study. You may also be asked to provide additional optional biofluid samples at the start or end of any specific treatments (e.g. at the start and end of radiotherapy).

We know that brain tumours can impact many areas of your life and we want to understand this better. To do this you will be asked to complete a 'Quality of Life' booklet that consists of four short questionnaires, initially when you join the study, then at key points in your treatment and when you are being monitored after treatment. These will generally be when you are attending the hospital to have your brain scans. A carer may assist you with completing the questionnaire, but the answers should be your own.

Patients joining BRAIN MATRIX will also be able to support a study, called BRIAN (the Brain tumouR Information and Analysis Network), run by the The Brain Tumour Charity.

BRIAN allows people to record their experiences of having a brain tumour using a mobile device or web app. This is to help doctors and researchers get a better understanding of how having a brain tumour affects the quality of life of patients who are living with a brain tumour or caring for someone who has a brain tumour. Your doctor will talk to you about participating in this additional study. In the future it is hoped that by analysing information collected by BRIAN together with all the other data collected from patients participating in BRAIN MATRIX, it will be possible to provide better overall care and support to patients and their families.

You can find out more about BRIAN and how to take part at the Brain Tumour Charity's website: https://www.thebraintumourcharity.org/living-with-a-brain-tumour/brian/.

You are welcome to take part in BRIAN even if you do not wish to take part in this study. Similarly, you can still participate in BRAIN MATRIX if you do not wish to join BRIAN.

#### How long will I be in the study?

We would like to continue to collect data on you for up to five years where possible.

#### What will I have to do?

If you decide to take part in this study, you will be required to:

- ✓ Sign a consent form to enter the study.
- ✓ Keep all scheduled medical and imaging appointments.
  (There are no additional appointments or scans as part of this study).
- ✓ At routine clinic visits your weight, neurological examination and performance status (measure of your general health and how your disease affects your daily routine) will be taken.
- ✓ Tell the study doctor about any medication that you are taking, even if it is medicine you buy without a prescription or is a natural or herbal remedy.
- ✓ Tell the doctor about any side effect, injury, symptom or complaint you experience, including any unplanned hospital admissions.
- ✓ Provide blood samples, and further optional biofluid samples, at the start and end of a treatment, and/or at tumour progression.
- ✓ Complete Quality of Life questionnaires when you join the study and when requested (usually at the start of a treatment, and/or at tumour progression, and otherwise every 3-6 months).

#### What are the alternatives for treatment?

If you choose not to take part in this study you will receive the same treatments as if you were on the study.

### 3. What are the possible benefits and risks of taking part?

#### What are the possible benefits of taking part?

We cannot promise that you will benefit directly from participating in this study. All the information that we get from this study will help improve the treatment of patients with glioma in the future.

By participating in this study, it will allow us to identify certain molecular characteristics of your tumour. These analyses can help refine and, in some cases, change the diagnosis. Your doctor will then be able to adapt and discuss the best treatment options for you.

This study may identify molecular changes in your tumour that suggest you could benefit from new-targeted treatments, which could be made available to you in the future or make you eligible for another clinical study/trial. Your doctor will discuss your options with you.

**Please note that identification of a target does not guarantee benefit from, or access to, a targeted treatment.**

#### What are the possible disadvantages and risks of taking part?

We want to try and improve the outcome for patients with glioma and believe that providing this standardised platform may improve outcomes in, and options for, patients. However, it is possible that this may not show any benefit over the current UK standard practice.

#### What about genetic results?

In addition to identifying changes in your tumour, it is possible that the genetic analysis might discover an unexpected genetic change in your normal cells. Genetic changes in normal cells may tell your doctors that you could be at risk of another disease. This might be a cancer or another disease that has nothing to do with your cancer. Changes like this could be present in other members of your family or passed on to your children. It is important to emphasise that the aim of this study is not to identify genes that may be inherited and cause cancer in families.

In the unlikely event that studies on your blood or tumour reveal genetic information that may affect you or other family members, we need to know whether you would wish to be informed. If you wish to be informed about these genetic changes, please initial the corresponding box on the informed consent form. We will then tell the doctor treating you at your local centre so that they can discuss the finding with you and arrange for any genetic counselling you may need. Please note that these unexpected genetic findings will not affect how your brain tumour is treated and will not prevent you from participating in this study.

You may choose to opt in or opt out of receiving information about genetic changes in normal cells on the informed consent form.

#### What are the side effects of the treatment?

There are no specific treatments within this study. Your doctor will discuss the side effects of your recommended treatment.

#### 4. What happens at the end of the study?

The study team will continue to collect data about your health and further treatment for up to 5 years where possible. This data will be collected at routine clinic appointments.

#### 5. Will anybody get paid if I take part?

You will not receive any money or study expense reimbursement for taking part in this clinical study.

#### 6. What if there is a problem?

If you have any concerns about your care during this study or any possible harm you may suffer, you should inform your study doctor immediately. More detailed information is given in Part 2.

#### 7. Will my taking part in this study be kept confidential?

Yes. We will follow ethical and legal practice and all information about your participation in this study will be kept confidential. The details of this are in Part 2 of this information sheet.

**This completes Part 1 of the Information Sheet**

If the information in Part 1 has interested you and you are considering participation, please read the additional information in Part 2 before making any decision.

### Part 2:
### What else do I need to know about taking part in this clinical study?

#### 8. More information about taking part

##### What if relevant new information becomes available?

Sometimes new information about the conditions being studied becomes available during the course of a research study. If this happens, your study doctor will tell you about it and discuss this with you. Your treatment is not affected by inclusion in this study, and therefore it is unlikely to influence whether you should continue in the study. If you decide not to carry on, your study doctor will make arrangements for your care to continue as normal. If you decide to continue with the study then you may be asked to sign an updated consent form. If the study is stopped for any other reason, your doctor will tell you and arrange your continuing care.

##### What if I decide that I don't want to carry on with the study?

You are free to withdraw from this study at any time. You do not have to give a reason for your decision and your future treatment will not be affected. Any data already collected prior to withdrawal may still be used for study purposes.

##### What if there is a problem?

In the event that something does go wrong and you are harmed because of taking part in the trial, and this is due to someone's negilence, then you may have grounds for claiming compensation from the Sponsor of the trial (the University of Birmingham) or the NHS Trust who treated you but you may have to pay your legal costs.

NHS Trust and non-Trust Hospitals have a duty of care to patients treated, whether or not the patient is taking part in a clinical trial, and the normal NHS complaints mechanism will still be available to you (if appropriate).

##### What will happen to any samples I give?

Collection of diagnostic biopsy samples, taken as part of your standard care, and blood samples at study entry are mandatory and are required for the genomic analysis to be performed. In addition, we will ask that pathology reports on your tissue samples are sent to the BRAIN MATRIX Study Office and to University of Oxford. Any samples remaining at the end of this analysis will be stored in a biobank and may be used in future ethically approved research, which may involve genetic analysis, animal or *in vitro* models, commercial or private institutions, and which may take place in the UK or overseas. If you change your mind about the use of your samples in the study, we will not collect any more blood samples and, if you request it, we will destroy all that remains of samples already taken.

##### Will my taking part in this study be kept confidential?

Yes. All information collected about you for this study will be subject to the General Data Protection Regulation (GDPR) and the Data Protection Act 2018 and will be kept strictly confidential. University of Birmingham is the Sponsor for this study based in the UK. We will be using information from your medical records in order to undertake this study and will act as the data controller for this study. This means that we are responsible for looking after your information and using it properly. University of Birmingham and the NHS will keep identifiable information about you for at least 25 years after the study has finished, allowing the results of the study to be verified if needed.

All information collected by the Sponsor will be securely stored at CRCTU at the University of Birmingham (BRAIN MATRIX Study Office) on paper and electronically and will only be accessible by authorised personnel. The only people at the University of Birmingham who will have access to information that identifies you will be people who manage the study or audit the data collection process. With your permission, your study doctor will provide your initials, date of birth, NHS/CHI number and hospital number when they enter you into the study and they will notify your GP that you intend to participate. They will also send a copy of your signed Informed Consent Form in the post to the BRAIN MATRIX Study Office.

The NHS will use your name and contact details to contact you about the research study, and make sure that relevant information about the study is recorded for your care, and to oversee the quality of the study. Your study doctor or research nurse may also need to send a copy of your Informed Consent Form to other healthcare professionals (e.g. your GP or NHS pathologist) to prove that you have given consent to take part in the study before they will provide information or tumour samples.

In the BRAIN MATRIX Study Office you will be identified by a unique study number. In routine communication between your hospital and the BRAIN MATRIX Study Office you will only be identified by your study number, initials and date of birth. We may need to record, and occasionally refer to you using your hospital, Genomics England (GEL) Referral ID number and histopathology numbers during analysis of your tumour samples and radiology numbers during analysis of your MRI scans. In addition, your study number, hospital, NHS, histopathology and radiology numbers may be included on samples sent to University of Oxford, University of Edinburgh and Genomics England for central review to help specialist pathologists and radiologists identify the tumour samples and MRI data.

All information will be treated as strictly confidential and nothing that might identify you will be revealed to any third party other than those involved in the treatment or organisation of tumour sample and imaging collection and transfer (e.g. staff at University of Birmingham, University of Oxford, University of Edinburgh and Genomics England). It may be necessary to send information about you such as study number and date of birth to the BRIAN databank (IRAS ID: 237931) run by The Brain Tumour Charity. This is for your, and others', protection to track the safety of the treatments used. We will also provide anonymised linked data to QMENTA, a neuroimaging storage platform, for research purposes. All third parties  have the same duty of confidentiality to you as all other research study personnel.

By taking part in the study you will be agreeing to allow research staff at your hospital, and from the BRAIN MATRIX Study Office, to look at the study records, and this includes your medical records. It may be necessary to allow authorised personnel from the University of Birmingham and/or NHS bodies to have access to your medical and research records. This is to ensure that the study is being conducted to the highest possible standards. We may also collect different aspects of your health data from the NHS and other Department of Health organisations, in addition to your medical records, for the purposes of long term followup.

From time to time we may be asked to share the study information (data) we have collected with researchers running other studies in this organisation and in other organisations, so that they can perform analysis on the data to answer other important questions about brain tumours. This may also reveal clinical trials or other research that you might benefit from, which your doctor or another member of your clinical team may discuss with you if appropriate. These organisations may be universities, NHS organisations or companies involved in health research and may be in this country or abroad. Any such request is carefully considered by the study researchers and will only be granted if the necessary procedures and approvals are in place. This information will not identify you and will not be combined with other information in a way that could identify you. The information will only be used for the purpose of health research, and cannot be used to contact you or to affect your care. It will not be used to make decisions about future services available to you, such as insurance. Under no circumstances will you be identified in any way in any report, presentation or publication arising from this or any other study.

All individuals who have access to your information have a duty of confidentiality to you.

If you choose to withdraw from the study treatment, we would still like to collect relevant information about your health, as this will be invaluable to our research. If you have any objection to this, please let your study doctor know.

You can withdraw your consent to our processing of your data at any time. Your rights to access, change, or move your information are limited, as we need to manage your information in specific ways in order for the research to be reliable and accurate. If you withdraw from the study, we will keep the information about you that we have already obtained. To safeguard your rights, we will use the minimum personally-identifiable information possible. Under the provisions of the General Data Protection Regulation you have the right to know what information the BRAIN MATRIX Study Office have recorded about you. If you wish to view this information or find more about how we use this information, please contact Legal Services at the address below.

Legal Services
University of Birmingham
Edgbaston
BIRMINGHAM
B15 2TT

More information about how your data will be handled can be found in the CRCTU Privacy Policy available on our website: www.birmingham.ac.uk/crctu.

#### Where can you find out more about how your information is used?

You can find out more about how we use your information in the GDPR Guidance document provided to you with this information sheet.

#### What will happen to the results of the study?

Results from the study will be published in medical journals but no individual patients will be identified. You will be able to get a copy of the published results by asking your doctor at the end of the study.

#### Who is organising and funding the research?

This research study is being carried out by a network of doctors across the UK. The study is sponsored by the University of Birmingham and co-ordinated by the Cancer Research UK Clinical Trials Unit at the University of Birmingham. The study is being funded by The Brain Tumour Charity, and some aspects of the genomic testing will be supported by Genomics England, part of Department of Health & Social Care. Your study doctor will not be paid extra for including you in this study.

##### Who has reviewed the study?

All research in the NHS is looked at by an independent group of people, called a Research Ethics Committee (REC) to protect your safety, rights, well-being and dignity. This study has been reviewed and given favourable ethical opinion by the West Midlands – Edgbaston Research Ethics Committee. It has also been approved by the local Trust Research and Development department at your hospital.

##### Involvement of the General Practitioner/Family Doctor (GP)

It is important that your GP is informed that you are taking part in this study. In addition, we may ask him/her to provide information on your progress. If we need to contact your GP for any follow-up information, we will need to use your full name in our correspondence.

#### 9. Who should I contact for further information?

##### Further information and contact details

If you have any questions or concerns about your disease or this clinical study, please discuss them with your doctor.

For more generic information about treating cancer, you may also find it helpful to contact '**About Cancer**', an information service about cancer and cancer research studies by Cancer Research UK:

**Freephone:** 0808 800 40 40

**Website:** www.cancerresearchuk.org/about-cancer/brain-tumours

For more information and support about brain tumours, you may find it helpful to contact **The Brain Tumour Charity**:

**Freephone:** 0808 800 0004

**Website:** www.thebraintumourcharity.org/

##### Your telephone contact numbers:

**Local Investigator:** ............................................. **Contact Number:** ...........................................

**Research Nurse:** ............................................. **Contact Number:** ...........................................

**Emergency (24 hours) Contact Number:** .......................................

**Thank you for taking time to read this Patient Information Sheet and considering taking part in this study.**

### Summary of Assessments

| Assessment / Activity | Screening | Platform Entry | Day 1 -28 | Start of concomitant/ adjuvant treatment | Further Surgery *(if applicable)* | Follow-up – up to 5 years |
|---|---|---|---|---|---|---|
| **Informed Consent** | ✓ | | | | | |
| **Confirm eligibility** | ✓ | | | | | |
| **Registration** | | ✓ | | | | |
| **Baseline Clinical Data Collection** | | ✓ | | | | |
| **Diagnostic Surgery** | | ✓ | | | ✓ | |
| **Collection of blood sample (mandatory)** | | ✓ | | | | |
| **Collection of blood samples (optional)** | | ✓ | | ✓ | ✓ | ✓ |
| **Neurological Examination** | | ✓ | | ✓ | ✓ | ✓ |
| **Performance Status** | | ✓ | | ✓ | ✓ | ✓ |
| **Weight** | | ✓ | | ✓ | ✓ | ✓ |
| **Quality of Life Questionnaires** | | ✓ | | ✓ | ✓ | ✓ |
| **Blood and biopsy samples sent to Oxford** | | ✓ | | | ✓ | |
| **Imaging data sent to Edinburgh** | | ✓ | | ✓ | ✓ | ✓ |
| **Local pathology report sent to BRAIN MATRIX Study Office and Oxford** | | | ✓ | | ✓ | |
| **Molecular testing and feedback** | | | ✓ | | ✓ | |
| **Reporting of relapse, further treatment** | | | ✓ | ✓ | ✓ | ✓ |
