## Supplementary Appendix 6 for "Protocol for the Tessa Jowell BRAIN MATRIX Platform Study"

### Supplementary Appendix 6: The Tessa Jowell BRAIN MATRIX Platform study schedule of events

| Activity* | Screening | Platform Entry | Day 1-28 | Initiation of concomitant therapy | Initiation of adjuvant therapy | Further surgery (if applicable) | Follow-up – up to 5 years# | Death |
|---|---|---|---|---|---|---|---|---|
| Informed consent[1] | X | | | | | | | |
| Confirm eligibility | X | | | | | | | |
| Registration | | X | | | | | | |
| Baseline Clinical Data Collection[2] | | X | | | | | | |
| Collection of blood for germline DNA | | X | | | | | | |
| Collection of Liquid biopsies – blood[3] | | X | | X | X | X | X | |
| Diagnostic surgery (biopsy or craniotomy) | | X | | | | X | | |
| Shipment of matched tumour and blood sample to Oxford BRAIN MATRIX Laboratory for molecular diagnosis[4] | | X | | | | X | | |
| Transfer of pseudo-anonymised imaging data to Edinburgh Imaging Hub[5] | | X | | X | X | X | X | |
| Clinician Global Impression of Change[6] | | | | X | X | X | X | |
| NIH Stroke Score[7] | | X | | X | X | X | X | |
| Weight[8] | | X | | X | X | X | X | |
| WHO Performance Status[9] | | X | | X | X | X | X | |
| Quality of Life questionnaires[10] | | X | | X | X | X | X | |
| Local pathology report sent to BRAIN MATRIX Study Office[11] | | | X | | | X | | |

| Activity* | Screening | Platform Entry | Day 1-28 | Initiation of concomitant therapy | Initiation of adjuvant therapy | Further surgery (if applicable) | Follow-up – up to 5 years# | Death |
|---|---|---|---|---|---|---|---|---|
| Reporting of relapse, further treatment and death | | | X | X | X | X | X | X |
| Post-mortem report and submit tissue specimens to CRUK PEACE study[12] | | | | | | | | X |

**Notes**

* Where applicable and acceptable in accordance to local practices, visits/assessments may be performed by telephone or video call.

### Follow-up visits to occur every 6 months and at clinic visit that coincides with the time point.

1. Written informed consent must be obtained within 28 days of Platform Entry and before any study-specific screening procedures.
2. Baseline Clinical Data Collection to include height.
3. Further optional blood samples should be collected whenever possible for future analysis of cell-free circulating tumour DNA (cfT-DNA) at the following time points: At first post-operative MRI; Initiation of adjuvant treatment (if applicable); the end of adjuvant treatment (if applicable); the time of objectively measured progression; during the palliative phase if a post-mortem has been agreed. Cerebrospinal fluid, if available for the patient, should also be submitted.
4. Local sites will register their samples (blood and frozen tissue) after the patient has been registered to the study and send them to the Oxford BRAIN MATRIX Laboratory for biobanking, biofluid analysis and complementary omics. For those cases that are recruited through the NHS Genomic Medicine Service (GMS) WGS pathway for Whole Genome Sequencing (WGS) in England;, a (small) tumour sample and one EDTA blood sample will be sent directly to the respective Genomics Laboratory Hub (GLH), once standard of care paired tumour blood WGS analysis is established. Patients must be consented to both the NHS GMS WGS pathway and BRAIN MATRIX. A copy of the Sample Form must also be sent to the BRAIN MATRIX Study Office. Please refer to the BRAIN MATRIX Platform Laboratory Manual for further details. Clinical data to also be included on the Sample Form.
5. Refer to the Imaging Manual for further details. Clinical data to also be included on Imaging Form. Neurosurgical navigation and radiotherapy planning imaging to be submitted in addition to MRI.
6. **Error! Reference source not found.**To be collected at the start and end of concomitant therapy and start and end of adjuvant treatment. For Further Surgery time point, to be collected at the patient's post-operative review. The Clinician Global Impression of Change score should also be collected around the time of imaging and submitted with the imaging to facilitate RANO assessment. The Clinician Global Impression of Change can be completed by Investigators or Research Nurses.
7. **Error! Reference source not found.**To be collected at the start and end of concomitant therapy and start and end of adjuvant treatment. For Further Surgery time point, to be collected at the patient's post-operative review. The NIH Stroke Score can be completed by Investigators or suitably trained Research Nurses.

8. Weight to be collected at the start and end of concomitant therapy and start and end of adjuvant treatment. For Further Surgery time point, to be collected at the patient's post-operative review.
9. **Error! Reference source not found.**To be collected at the start and end of concomitant therapy and start and end of adjuvant treatment. For Further Surgery time point, to be collected at the patient's post-operative review.
10. QoL Questionnaires to be completed at follow-up visits that coincide with imaging appointments. **Error! Reference source not found.**
11. Pseudonymised copy of local pathology report for each sample must be sent to the BRAIN MATRIX Study Office as soon as it is available. Once received, it will be shared with the Oxford BRAIN MATRIX Laboratory for Genomic Tumour Advisory Board discussion. Submission of this report is the responsibility of the recruiting site.
12. Separate consent to the CRUK PEACE study (or equivalent) to be obtained.
