## Supplementary Appendix 7 for "Protocol for the Tessa Jowell BRAIN MATRIX Platform Study"

### Supplementary Appendix 7: Definition of adverse events

#### Adverse Event (AE)

Any untoward medical occurrence in a patient or clinical trial subject participating in the study which does not necessarily have a causal relationship with the treatment received.

Comment:

An AE can therefore be any unfavourable and unintended sign (including abnormal laboratory findings), symptom or disease temporally associated with the use of a medicinal product, whether or not related to the medicinal product.

#### Related Event

An event which resulted from the administration of any of the research procedures.

#### Serious Adverse Event (SAE)

An untoward occurrence that:

- Results in death
- Is life-threatening*
- Requires hospitalisation** or prolongation of existing hospitalisation
- Results in persistent or significant disability or incapacity
- Consists of a congenital anomaly/ birth defect
- Or is otherwise considered medically significant by the Investigator***

Comments:

The term severe is often used to describe the intensity (severity) of a specific event. This is not the same as serious, which is based on patients/event outcome or action criteria.

* Life threatening in the definition of an SAE refers to an event in which the patient was at risk of death at the time of the event; it does not refer to an event that hypothetically might have caused death if it were more severe.

** Hospitalisation is defined as an unplanned, formal inpatient admission, even if the hospitalisation is a precautionary measure for continued observation. Thus, hospitalisation for protocol treatment (e.g., line insertion), elective procedures (unless brought forward because of worsening symptoms) or for social reasons (e.g., respite care) are not regarded as an SAE.

*** Medical judgment should be exercised in deciding whether an AE is serious in other situations. Important AEs that are not immediately life threatening or do not result in death or hospitalisation but may jeopardise the subject or may require intervention to prevent one of the other outcomes listed in the definition above, should be considered serious.

##### Unexpected and Related Event

An event which meets the definition of both an Unexpected Event and a Related Event.

##### Unexpected Event

The type of event that is not listed in the protocol as an expected occurrence.
